# Nitrate reduction capacity of the oral microbiota is impaired in periodontitis: potential implications for systemic nitric oxide availability

**DOI:** 10.1101/2023.06.21.23291703

**Authors:** Bob Rosier, William Johnston, Miguel Carda-Diéguez, Annabel Simpson, Elena Cabello-Yeves, Krystyna Piela, Robert Reilly, Alejandro Artacho, Chris Easton, Mia Burleigh, Shauna Culshaw, Alex Mira

**Affiliations:** Department of Genomics and Health, FISABIO Foundation, Center for Advanced Research in Public Health, Valencia, Spain; Department of Biological and Biomedical Sciences, Glasgow Caledonian University, Glasgow, UK; Oral Sciences, University of Glasgow Dental School, School of Medicine, Dentistry and Nursing, College of Medical, Veterinary and Life Sciences, University of Glasgow, UK; Sport and Physical Activity Research Institute, University of the West of Scotland, Blantyre, Scotland; Instituto de Biomedicina de Valencia, Consejo Superior de Investigaciones Científicas (IBV-CSIC), Valencia, Spain; CIBER Center for Epidemiology and Public Health, Madrid, Spain

## Abstract

**Background:** The reduction of nitrate to nitrite by the oral microbiota has been proposed to be important for oral health and results in nitric oxide formation that can improve cardiometabolic conditions, such as hypertension and diabetes. Studies of bacterial composition in subgingival plaque suggest that nitrate-reducing bacteria are associated with healthy conditions, but the impact of periodontitis on nitrate-reducing capacity and therefore nitric oxide availability has not been evaluated. The aim of the current study is to see assess how periodontitis and periodontal treatment affect the nitrate reduction capacity of the oral microbiota.

**Methods:** First, 16S rRNA sequencing data of five studies from different countries were analysed with the Dada2 pipeline to compare nitrate-reducing bacteria in health and periodontitis. Additionally, subgingival plaque, saliva and plasma samples from 42 periodontitis patients were collected before and after non-surgical periodontal therapy (NSPT). The subgingival plaque bacterial composition was determined using Illumina sequencing of the 16S rRNA gene and the quantity of the nitrate-reducing biomarker genus *Rothia* was determined by qPCR. Measurements of nitrate and nitrite in saliva and plasma were performed and the salivary nitrate reduction capacity (NRC) was determined after three hours of incubation *in vitro* and compared with the NRC of 15 healthy individuals.

**Results:** Nitrate-reducing bacteria were significantly lower in subgingival plaque of periodontitis patients compared with healthy individuals (p < 0.05 in all five datasets). After NSPT, nitrate-reducing bacteria increased in subgingival plaque (p < 0.05) and correlated negatively with periodontitis-associated bacteria (p < 0.001). A post-treatment increase in the genus *Rothia* was confirmed by qPCR (p < 0.05), as well as an increase in the salivary NRC (p < 0.05). No significant effect was found of NSPT on the plasma nitrate and nitrite levels in this population.

**Conclusion:** The levels of nitrate-reducing bacteria of the subgingival microbiota decrease in periodontitis and both their levels and the salivary NRC increase after NSPT. We hypothesize that an impaired NRC can affect nitric oxide availability and can therefore be an instrumental link between periodontitis and systemic conditions.

## Introduction

The accumulation of dental plaque due to insufficient oral hygiene can facilitate the development of gingivitis ^1^. Gingivitis is mostly reversible, but long or repeated episodes of gingivitis, especially in susceptible individuals, can lead to the development of periodontitis - a chronic and destructive inflammatory disease in which host tissue is lost. In periodontitis, periodontal pockets develop in which the subgingival plaque microbiota shifts towards a disease-associated composition, including an increase of anaerobic, proteolytic, inflammation-tolerant and/or alkalophilic species ^2–4^.

Along with an increase in disease-associated bacteria, a decrease in health-associated bacteria has also been observed ^5, 6^. A healthy subgingival environment has normally been associated with the dominance of aerobic or facultatively anaerobic organisms ^3^. However, closer investigation of these healthy microbial populations reveal that they include all genera that up-to-date have been confirmed to reduce nitrate by physiological measurements, namely *Rothia*, *Neisseria*, *Actinomyces*, *Veillonella*, *Kingella*, and *Propionibacterium* ^7^. Of these genera, *Rothia* and *Neisseria* are the bacteria with the strongest association with nitrate, increasing in most (if not all) studies in which oral communities are exposed to nitrate ^8, 9^. It has been estimated that we obtain over 80% of dietary nitrate from vegetables – a food group strongly associated with systemic health benefits. However, the relationship between periodontitis, nitrate-rich foods and systemic health consequences has yet to be elucidated.

For centuries, it has been known that the impact of periodontitis is not isolated to the oral cavity, where it can lead to inflammation, bleeding, halitosis, and tooth loss. The oral cavity is the beginning of the respiratory system and gastrointestinal tract and directly connected to the blood stream via highly vascularized oral tissues ^10^. Periodontitis is associated with an increased risk of diabetes, rheumatoid arthritis, atherosclerosis, hypertension, pregnancy complications, and Alzheimer’s disease, among others ^11^. This periodontal-systemic link gave rise to the concept of periodontal medicine ^12^, which from a mechanistic point of view, has been explained as the effect of periodontitis-associated bacteria, their products and/or inflammatory molecules produced in the inflamed gingiva reaching different parts of the body and causing complications through a systemic inflammation state ^10, 11^.

In contrast to an increase in disease-associated species causing systemic complications, some oral microbiota products can result in health benefits. For example, nitrate-reducing bacteria in the mouth reduce nitrate into nitrite and, in some cases, further to nitric oxide ^13, 14^. The nitric oxide produced by oral bacteria can enter the blood stream directly via the oral mucosa, whilstpart of the salivary nitrite is swallowed and converted into nitric oxide in the stomach (by acidic decomposition of nitrite) and host tissue (e.g., by the reaction of nitrite with hemin) ^15^. This can have several cardio-metabolic benefits, including lowering of blood pressure, improved endothelial function, greater efficiency of exercise performance, and reversal of metabolic syndrome, as well as antidiabetic effects ^16, 17^. Therefore, it is crucial to evaluate if, in addition to the pro-inflammatory effect of periodontal pathogens, a lowered capacity to utilize dietary nitrite could be taking place during periodontitis that could also contribute to the systemic effects of this disease.

In addition to systemic benefits, nitrate metabolism by the oral microbiota appears to be important for oral health (reviewed by Rosier et al., ^7^). Nitric oxide has antimicrobial properties, killing sensitives species, including anaerobic bacteria associated with periodontitis ^18^. In contrast, representatives of *Rothia* and *Neisseria* increase in the presence of nitrate and are associated with the absence of inflammation. Nitric oxide could also directly signal to epithelial cells, possibly contributing to gingival homeostasis by increasing blood flow and mucus thickness, while decreasing inflammation.

In the current work, we therefore aim to evaluate if the levels of nitrate-reducing bacteria decrease in periodontitis, as well as if the nitrate-reduction capacity in periodontal patients is impaired, in order to assess if this could contribute to the link between periodontitis and systemic complications. Additionally, we also aim to determine if nitrate-reducing genera and nitrate reduction capacity increase after non-surgical periodontal therapy (NSPT). For this, 16S rRNA sequencing data of subgingival samples were analysed to determine if nitrite-producing bacteria were lower in periodontitis compared to periodontal health. Additionally, subgingival plaque, saliva and plasma samples were collected from periodontitis patients before and after non-surgical periodontal therapy (NSPT). Nitrate and nitrite were measured in saliva and plasma and the salivary nitrate reduction capacity (NRC) was determined by incubating saliva with a physiological nitrate concentration. To examine the effect of NSPT on nitrite-producing bacteria in subgingival plaque, the overall bacterial composition was determined using Illumina sequencing and the genus *Rothia*, considered a consistent biomarker of nitrate reduction, was measured by qPCR.

## Materials and Methods

### Bioinformatic analysis comparing health and periodontitis

A bioinformatic analysis of previously published datasets was performed to find differences in the nitrite-producing microbiota between periodontal health and periodontitis (Figure 1A). Datasets containing 16S rRNA sequencing data from subgingival plaque samples were downloaded from the NCBI SRA Database and included individuals from Japan ^19^, Brazil ^20^, Chile ^21^, USA ^22^ and Spain ^23^. The Fastq files were processed as previously described using DADA2 R Statistics package (v1.20.0) ^24, 25^. Briefly, R1 and R2 reads were trimmed by length, and reads with more than 5 errors were removed. Reads were dereplicated to obtain true sequence variants that were then merged (min overlap 15 bp) and annotated to the SILVA v.138.1 database ^26, 27^. Bacteria were classified as nitrite-producers or confirmed nitrate reducers based on Rosier et al. ^7^. Additionally, species were classified as periodontitis-associated, including the red and orange complexes identified by Socransky et al. ^28^ and periodontitis-associated bacteria identified by Perez-Chaparo et al. ^29^ or just red complex. The exact list of species in the four groups (nitrite producers, confirmed nitrate reducers, periodontitis-associated and red complex) can be found in Supplementary Table 1.

**Figure 1:**
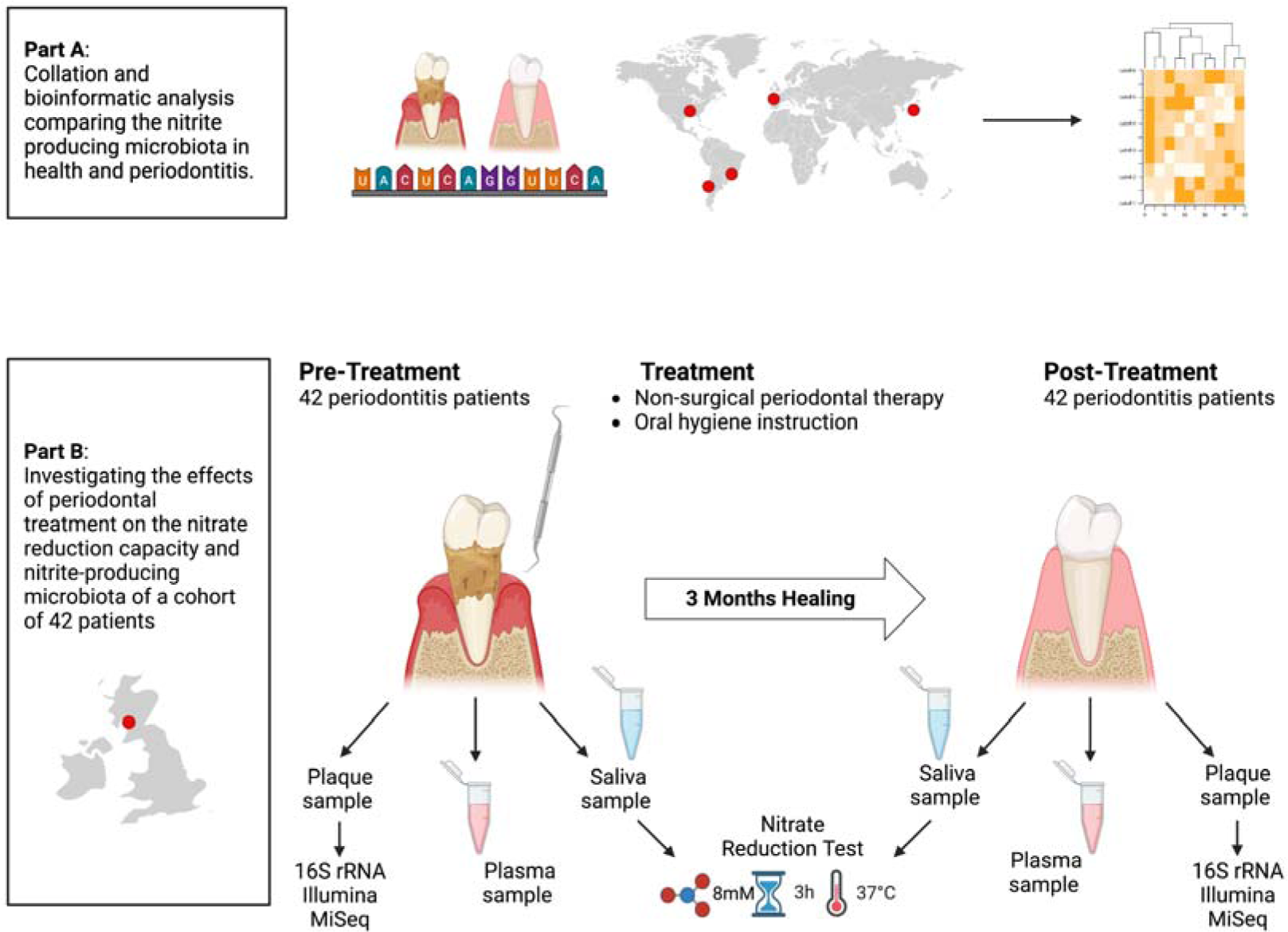
study overview. A) 16S rRNA sequencing data of five studies from five different countries (Japan, Brazil, Chile, USA and Spain) were analysed with the Dada2 pipeline to compare nitrate-reducing bacteria in health and periodontitis. B) Subgingival plaque, saliva and plasma samples from 42 periodontitis patients located in Glasgow (Scotland) were collected before and after non-surgical periodontal therapy (NSPT). The subgingival plaque bacterial composition was determined using Illumina sequencing of the 16S rRNA gene and the quantity of the nitrate-reducing biomarker genus Rothia was determined by qPCR. Measurements of nitrate and nitrite in saliva and plasma were performed and the salivary nitrate reduction capacity (NRC) was determined after three hours of incubation in vitro and compared with the NRC of 15 healthy individuals.

### Bacterial composition in periodontitis patients before and after periodontal treatment

To study the effect of periodontal treatment on the nitrate-reducing microbiota, data and samples were used of study previously described in Davison et al. ^30^ and Johnston et al. ^31^. The study was conducted in accordance with the Declaration of Helsinki (2013) and received ethical approval (London-Stanmore Research Ethics Committee, Reference: 14/LO/2064). The patients were recruited at the Glasgow Dental Hospital and periodontitis was defined as probing pocket depths ≥ 5 mm on 2 or more teeth at non-adjacent sites excluding third molars. All patients signed a written informed consent prior to participation. Other inclusion criteria and sample collection are described in Johnston et al. (2021). In short, 42 patients were included and received non-surgical periodontal treatment (NSPT) by a single experienced dental hygienist. Subgingival plaque (collected with a curette by a single experienced dental hygienist), drooling saliva and plasma samples were taken before any treatment visits (baseline) and 90-days following the last treatment visit (day 90). At day 90, periodontal parameters had improved significantly ^31^. Additionally, healthy control saliva samples were collected from volunteers under the study “Host-microbiota interactions in oral health and disease”, project number: 2011002. The healthy control study received ethical approval from the University of Glasgow MVLS ethical committee. All samples were stored at −80°C prior to usage.

DNA was extracted from subgingival samples (baseline and day 90) using the MagNA Pure LC DNA isolation kit (Roche Diagnostics, Mannheim, Germany) with the addition of a chemical lysis step with an enzymatic cocktail containing lysozyme, mutanolysin and lysostaphin, following Rosier et al. ^9^. As previously described by Johnston et al. ^31^, DNA concentrations were measured using a QubitTM 3 Fluorometer (Thermofisher, Waltham, Massachusetts, USA). An Illumina amplicon library was prepared following the 16S rRNA gene Metagenomic Sequencing library preparation Illumina protocol (Part #15,044,223 Rev. A). The primer sequences used in this protocol were; Illumina_16S_341F (TCGTCGGCAGCGTCAGATGTGTATAAGAGACAGCCTACGGGNGGCWGCAG) and Illumina_16S_805R (GTCTCGTGGGCTCGGAGATGTGTATAAGAGACAGGACTACHVGGGTATCTAATCC) which target the 16S V3 and V4 region. Following amplification, DNA was sequenced with an Illumina MiSeq Sequencer according to manufacturer’s instructions using the 2lll×lll300 base paired-ends protocol. For taxonomic classification, an amplicon sequence variant (ASV) table was obtained using the DADA2 pipeline in R ^24^. Taxonomy was assigned by comparison to the SILVA database ^26^, where the naive Bayesian classifier was used to assign sequences at the species-level. Bacterial species were classified as nitrite-producing or periodontitis-associated (Supplementary Table 1).

#### qPCR of Rothia in subgingival plaque

The total amounts of *Rothia* cells in subgingival plaque were analysed through quantitative PCR (qPCR) amplification of the *Rothia* nitrate reductase *narG* gene as described by Rosier et al. ^32^. Primers sequences were designed to be specific for the *Rothia* genus, using conserved regions of narG from *Rothia mucilaginosa*, *R. dentocariosa* and *R. aeria*. The forward primer sequence was 5’-ACA CCA TYA AGT ACT ACGG-3’ and the reverse 5’-TAC CAG TCG TAG AAG CTG-3’. Reactions of 20 μl were added per well of a qPCR plate, consisting of 10 μl of Light Cycler 480 SYBR Green I Master mix (Roche Life Science, Penzberg, Germany), 0.4 μl of each specific primer (10 μM), 8.2 μl water and 1 μl of template DNA (DNA isolated from subgingival plaque samples). Each sample was added in duplicate and measurements were performed using a Light Cycler 480 Real-Time PCR System (Roche Life Science) with the following conditions: 95°C for 2 min, and 40 cycles of 95°C for 30 s, 60°C for 20 s, and 72°C for 25 s. Negative controls were added, as well as a standard curve, consisting of a series dilution of an equimolar DNA mix of three *Rothia* species (*R. mucilaginosa* DSM-20746, *R. dentocariosa* DSM-43762, *R. aeria* DSM-14556) quantified with a QubitTM 3 Fluorometer (Thermofisher). Based on genome sizes ^33^, the number of *Rothia* cells was calculated, assuming a single copy of the *narG* gene per cell. The samples of five individuals out of 42 individuals had no DNA left after the sequencing procedure, leading to qPCR data of 37 individuals.

### Salivary nitrate reduction test

The nitrate reduction capacity (NRC) in periodontal patients (n=42) and healthy controls (n=15) was determined by defrosting saliva samples on ice and incubating this for 3 hours at 37°C in the presence of 8 mM nitrate. For this, 25 µl of water with 80 mM sodium nitrate (Sigma) was added to 225 µl of saliva in an Eppendorf tube.

#### Nitrate and nitrite in saliva

For nitrate and nitrite measurements in saliva, the RQflex 10 Reflectoquant (Merck Millipore, Burlington, Massachusetts, USA) reflectometer was used as described by Rosier et al. ^9^. The test strips (Reflectoquant, Merck Millipore) for nitrate had a range of 3–90 mg/L and the strips for nitrite a range of 0.5–25 mg/L. Accuracy of both reflectometer methods was confirmed using standard solutions (Merck Millipore) with known concentrations of the different compounds. The saliva was used directly or diluted 5-10 fold depending on the nitrate and nitrite concentrations. Fifteen µL of (diluted) saliva was added to each of the two reactive patches on a strip and excess liquid was removed by tipping the side of the strip on a tissue.

Before nitrate measurements, the diluted supernatants were treated with amidosulfuric acid (Sigma-Aldrich) based on the manufacturer’s instructions. For this, 35 µL of diluted supernatant was mixed with 1.5 µL amidosulfuric acid solution (10%).

#### Nitrate and nitrite measurements in plasma

Plasma nitrate and nitrite levels were determined using ozone-based chemiluminescence as described by Liddle et al. ^34^. For the measurement of plasma nitrite, tri-iodide reagent and 100 μL of anti-foaming agent were placed into a purge vessel which was heated to 50 °C. A standard curve was produced by injecting 100 μL of nitrite solutions (62.5-1000 nM) and a control sample (0 nM). Following this, plasma samples were thawed in a water bath at 37 °C for 3 min and 100 μL of the sample was injected into the purge vessel in duplicate. The concentration of NO cleaved during the reaction was then measured by the NO analyser (Sievers NOA 280i, Analytix, UK). For the measurement of plasma nitrate, vanadium reagent and 100 μL of anti-foaming agent were placed into the glass purge vessel and heated to 95 °C. A standard curve was produced by injecting 25–50 μL of nitrate solutions (6.25-100 μM) and a control sample (0 μM). Plasma samples were thawed and de-proteinised. Subsequently, 50 μL of the sample was injected into the purge vessel in duplicate and plasma nitrate calculated as previously described for the nitrite assay.

### Statistical analysis

Statistical analysis of nitrate and nitrite in saliva and plasma and *Rothia* cells in subgingival plaque (determined by qPCR) were performed using a nonparametric Wilcoxon test using IBM SPSS statistics (version 27) or GraphPad (version 9.5.1) and considered statistically significant at p-value < 0.05.

For analysis of the bacterial groups (i.e., nitrite producers, confirmed nitrate reducers, periodontitis-associated and red complex), R programming language (v3.4lll+) was used for statistical computing. The abundance of species was standardized using ANCOM-BC ^35^. The sums of standardized compositional data of bacteria in different groups were then compared using the Wilcoxon signed rank tests (wilcox.test function of stats library of R) and considered statistically significant at adjusted p-value < 0.05.

Correlations within and between the relative abundances of groups of bacteria and physiological parameters were determined with Spearman’s rho (cor.test function of stats library of R), along with associated adjusted p-value. Apart from the parameters of this study, correlations with salivary cytokines [tumour necrosis factor α (TNFα), interleukin-6 (IL-6) and interleukin-1β (IL-1β)] and clinical parameters [Periodontal pocket depths (PPD), clinical attachment level (CAL), full mouth bleeding score (FMBS), full mouth plaque score (FMPS) and periodontal inflamed surface area (PISA)] obtained by Johnston et al. ^31^ were explored.

Figures 1 and 2 and Supplementary Figure 1 were created in Microsoft Excel and/or using BioRender, and all other Figures were assembled using GraphPad PRISM (version 9.5.1).

**Figure 2:**
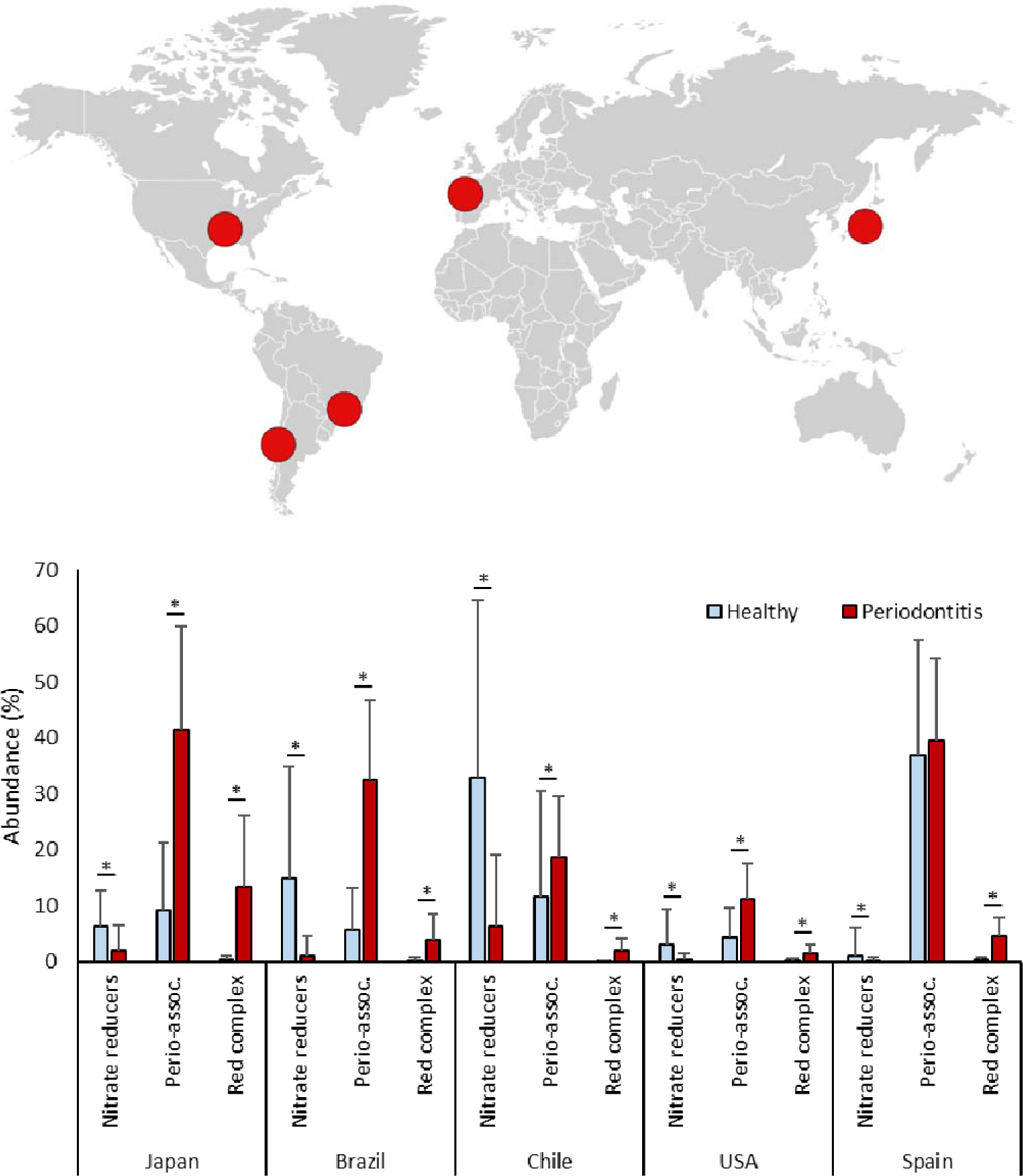
Confirmed nitrate-reducing bacteria in periodontitis and health. Bar graphs show the relative abundance of bacteria in subgingival plaque samples from different countries, as estimated by high-throughput sequencing of the 16S rRNA gene. Bacteria were grouped in confirmed nitrate-reducing species, periodontitis-associated or “red-complex” periodontal pathogens according to Rosier et al., Pérez-Chaparro et al. and Socransky et al. respectively (the bacterial species in each group are listed in Supplementary Table 1). In Supplementary Figure 1, all known nitrite-producing bacteria (some which may produce nitrite by other pathways than nitrate reduction) are shown following the same pattern and significant difference between health and periodontitis. The datasets include individuals from Japan (n = 10 periodontitis patients and 10 healthy individuals), Spain (n = 22 periodontitis patients and 60 healthy individuals), USA (n = 29 periodontitis patients and 28 healthy individuals), Brazil (n = 27 periodontitis patients and 21 healthy individuals) and Chile (n = 22 periodontitis patients and 17 healthy individuals). *adjusted p < 0.05 of compositional data standardized by ANCOM-BC and compared with a Wilcoxon test.

## Results

### Levels of nitrate-reducing bacteria in periodontal patients

A total of five datasets were analyzed where 16S rRNA gene sequencing of subgingival samples had been performed, spanning individuals in Japan (n = 10 periodontitis patients and 10 healthy individuals), Spain (n = 22 periodontitis patients and 60 healthy individuals), USA (n = 29 periodontitis patients and 28 healthy individuals), Brazil (n = 27 periodontitis patients and 21 healthy individuals) and Chile (n = 22 periodontitis patients and 17 healthy individuals). As expected, a higher proportion of “Red complex” bacteria directly involved in periodontitis was found in periodontal patients compared to healthy individuals in all five countries (Figure 2). In relation to the denitrifying microbiota, the relative proportion of nitrate-reducing bacteria varied between datasets. Regardless of the levels in healthy individuals in each country, the proportion of nitrate-reducing bacteria (Figure 2) were significantly lower in periodontal patients in all cases (p < 0.05 in all countries). When considering all known nitrite-producing bacteria (some which may produce nitrite by other pathways than nitrate reduction), the pattern and statistical significance remained (Supplementary Figure 1).

### Levels of nitrate-reducing bacteria before and after periodontal treatment

The bacterial composition of subgingival samples before and after a NSPT showed a consistent pattern across individuals. Relative to baseline, there was a significant increase in bacteria capable of reducing nitrate (Figure 3A, Supplementary Figure 2), with an opposite pattern for periodontal pathogens, including both the three species of the “red complex” and the extended list of periodontitis-associated bacteria, as reported previously by Johnston et al. ^31^. This suggests that the capacity to metabolize nitrate by subgingival plaque is improved after NSPT. The increase in the levels of nitrate reducing bacteria was confirmed by qPCR using specific primers for the nitrate-reducing biomarker *Rothia*, which showed a significant increase in the absolute levels (Figure 3B) and a trend of increase of the normalized levels of this organism (Figure 3C). Both the absolute and normalized levels of *Rothia* determined with qPCR correlated with the abundance of nitrate-reducing bacteria (r = 0.592 and r = 0.574, respectively, both p < 0.001 when grouping BL and D90) (Supplementary Datasheet).

**Figure 3:**
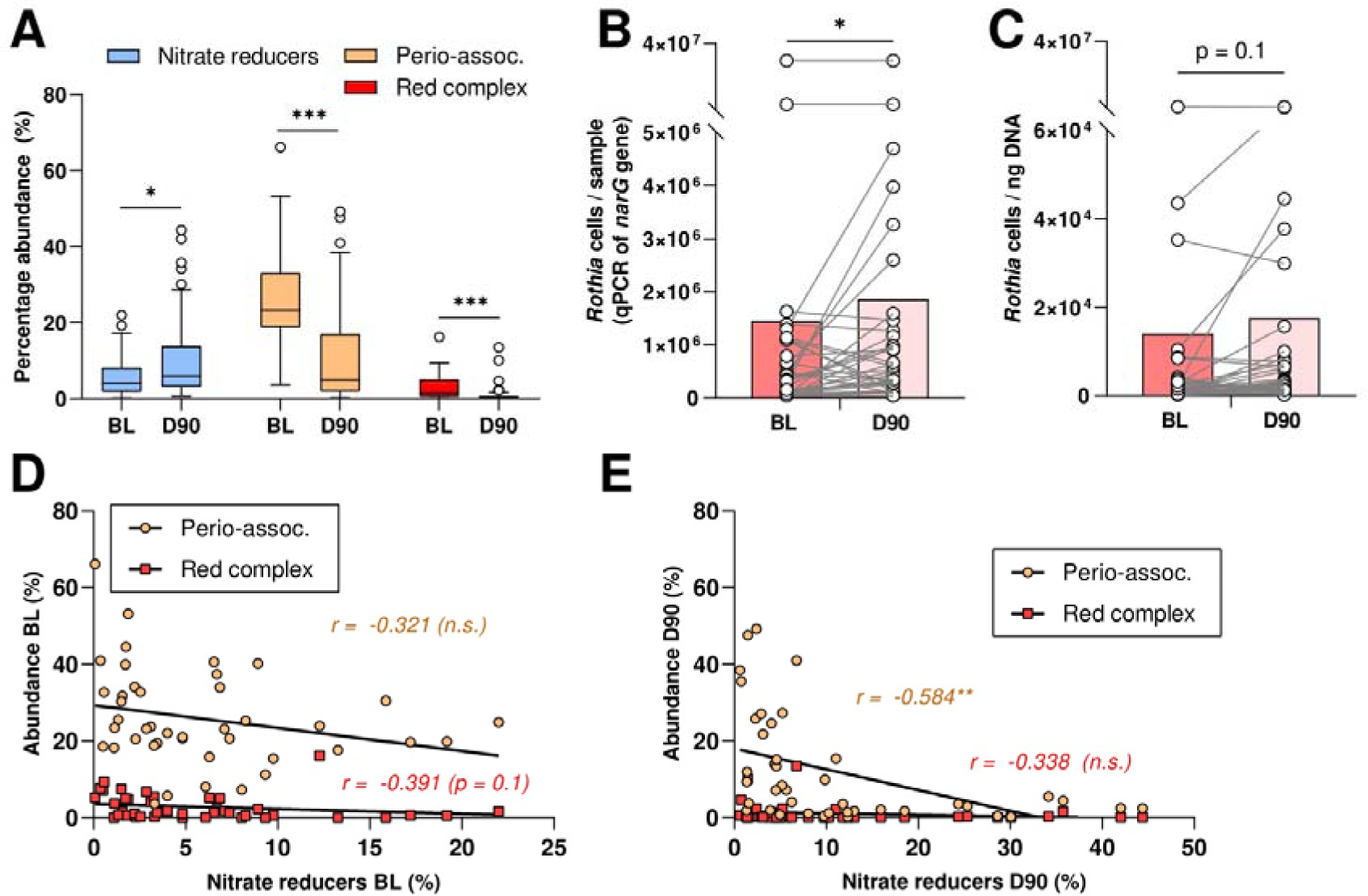
Confirmed nitrate-reducing species and disease-associated bacteria before and 90 days after periodontal treatment. A) Relative abundances of confirmed nitrate-reducing bacteria, red complex and periodontitis-associated bacteria before (baseline, BL) and 90 days after treatment (D90) of 42 periodontitis patients. *adjusted p < 0.05, *** p < 0.001 of compositional data standardized by ANCOM-BC and compared with a Wilcoxon test. B and C) Rothia cells determined by qPCR before (BL) and 90 days after (D90) treatment per sample (absolute amount, n = 37) (B) or per ng of DNA (C). *p < 0.05 determined by a Wilcoxon test. D and E) Correlations between abundance of periodontal pathogens and nitrite-producing bacteria at baseline (BL) and 90 days after treatment (D90). **adjusted p < 0.01 of Spearman’s rank correlation (n.s. = not significant). In Supplementary Figure 2, the comparisons and correlations of all known nitrite-producing bacteria (some which may produce nitrite by other pathways than nitrate reduction) are shown.

A significant negative correlation was found between the levels of nitrate reducing bacteria and the levels of periodontitis-associated bacteria (r = −0.523, p < 0.001 when grouping BL and D90) (Supplementary Datasheet). Before treatment (BL), there was a trend of negative correlation between nitrate-reducing bacteria and the red complex and after treatment (D90), there was a clear negative correlation between nitrate-reducing bacteria and periodontitis-associated bacteria (Figure 3D and 3E). The group of all nitrite-producing bacteria (Supplementary Table 1) correlated negatively with salivary IL-1β at baseline (r = −0.547, p < 0.01) (Supplementary Datasheet).

### Nitrate reduction capacity in health and periodontitis

Nitrate utilization during the incubation of saliva samples in the presence of nitrate for a period of three hours was considered an estimate of the nitrate reduction capacity (NRC) in each individual. The data revealed that nitrate reduction was higher in healthy individuals compared to periodontal patients at baseline, where nitrate levels did not change over the three-hour incubation period (Figure 4A). In accordance with the 16S rRNA sequencing data, the NRC in periodontal patients was restored after the periodontal treatment, with a significant depletion in nitrate levels after the incubation. This confirms that periodontal treatment induces a rise in the levels of nitrate-reducing bacteria in subgingival plaque and in the capacity to metabolize nitrate.

**Figure 4:**
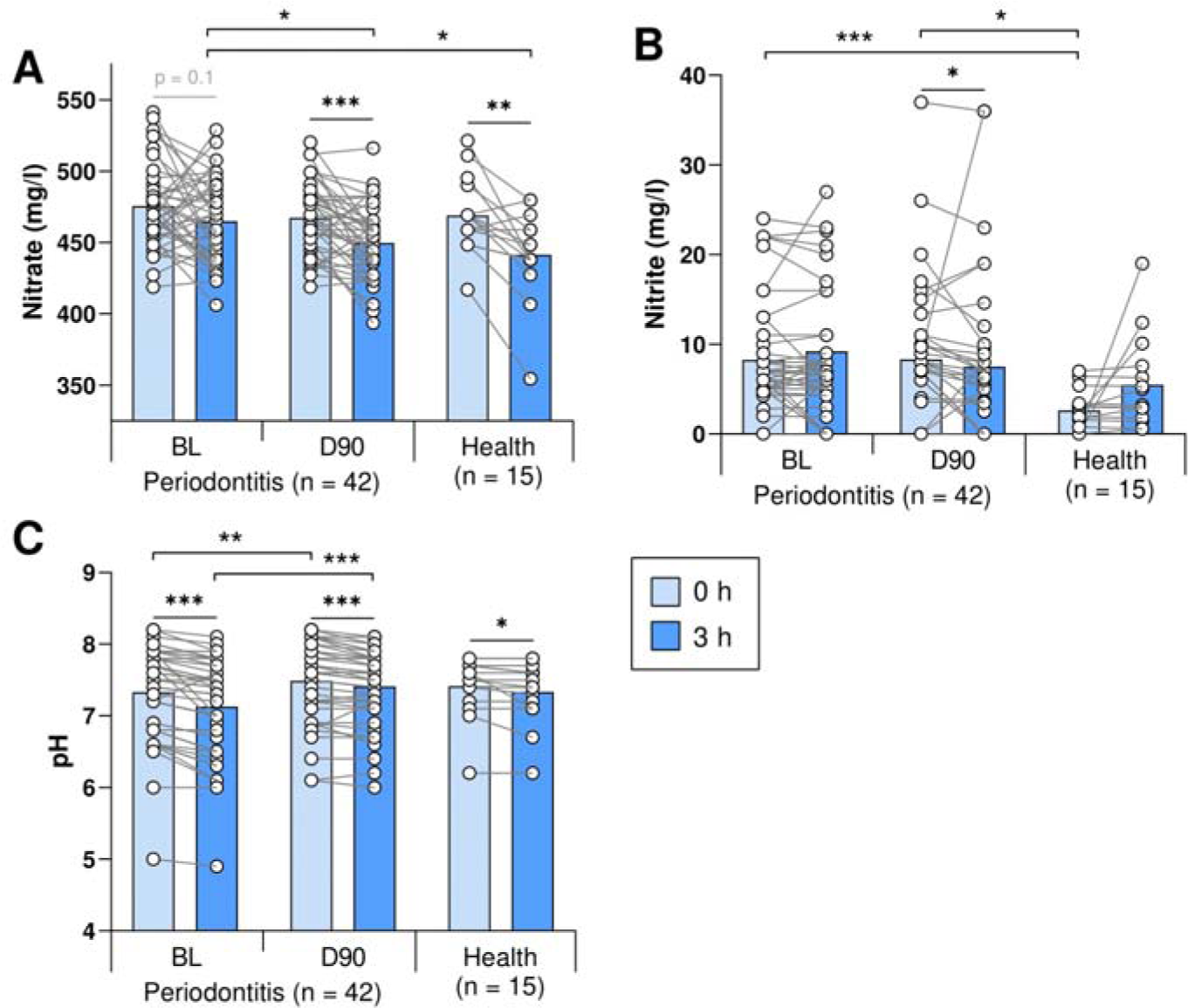
Nitrate reduction capacity (NRC) under health and periodontitis. A and B) Bar graphs represent the concentration of nitrate (NO^3^ _-_) and nitrite (NO^2^ _-_) of saliva samples cultured in vitro for 3 hours in the presence of 8 mM nitrate at 37°C before (Baseline, BL) and 90 days after periodontal treatment (N = 42) of periodontal patients, compared to healthy controls (n = 15). C) Bars show the pH during this incubation period for the same groups of individuals. Asterisks represent statistically significant differences (*p < 0.05, **p < 0.01, ***p < 0.001).

The activity of nitrate-reducing bacteria was also confirmed indirectly by pH measurements, which showed a larger pH drop in periodontal patients at baseline than after treatment (Figure 4C). This is expected, as nitrate reduction is known to buffer pH drops by proton consumption during denitrification or nitrite reduction to ammonium ^7^.

In agreement with the above, nitrite concentration levels did not differ in baseline periodontal samples before and after the incubation with nitrate (Figure 4B), confirming a hampered nitrate-reducing capacity. After periodontal treatment, a significant decrease in nitrite was observed after the incubation. Given the nitrate consumption during this three-hour period, the lower nitrite levels are probably a consequence of further nitrite reduction into nitric oxide ^13^. In healthy individuals, nitrite levels did increase after the incubation, but this increase was not significant (p = 0.17) and a much lower initial concentration of nitrite was present compared with periodontitis before (p < 0.001) and after (p < 0.05) treatment.

### Plasma levels of nitrate and nitrite

Contrary to the salivary levels of nitrate and nitrite, quantification of nitrate and nitrite concentration in morning blood samples collected in periodontal patients at baseline and 90 days after the periodontal treatment revealed no statistical differences between these two time points (Figure 5A-B). Plasma nitrate levels at both time points were on average around 50 uM, which represents a concentration about 10 times lower the physiological blood concentration after a nitrate-rich meal ^15^. Nitrite levels were over 400 times lower than nitrate (Figure 5C), indicating that at the timing that the samples were collected, nitrate-reduction metabolism was not detected. No samples were collected after nitrate supplementation. There was a trend of correlation between absolute and normalized *Rothia* levels in subgingival plaque and plasma nitrite (r = 0.278 and 0.278, respectively, both p = 0.1 when grouping BL and D90) (Supplementary Datasheet).

**Figure 5:**
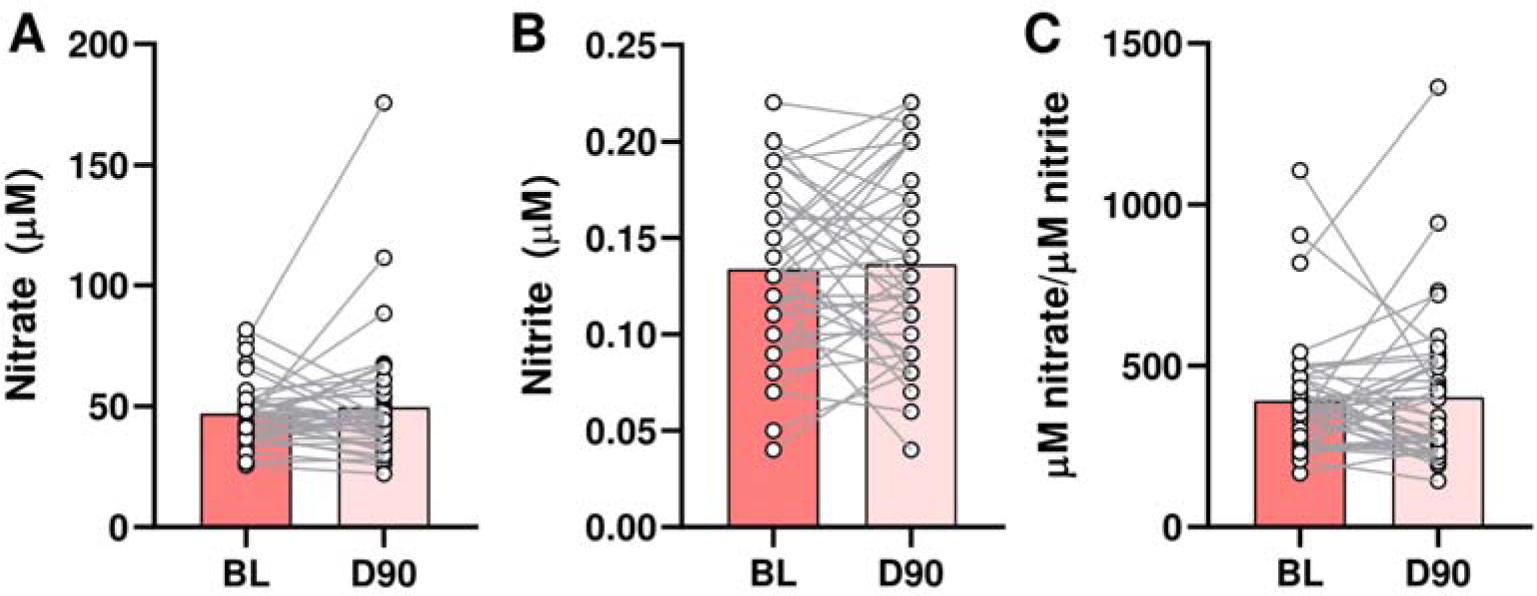
Plasma nitrate and nitrite before and 90 days after periodontal treatment. Data is shown of 42 periodontitis patients before (BL) and after (D90) treatment. A) Plasma nitrite. B) Plasma nitrate. C) Ratio nitrate / nitrite.

## Discussion

In this study, we used 16S sequencing datasets of 5 different countries (Japan, Brazil, Chile, USA and Spain) to compare the subgingival plaque microbiota in periodontitis and health. All countries followed the same pattern of a decrease in periodontitis-associated species and an increase in nitrite-producing bacteria, including confirmed nitrate-reducing species. This trend is in agreement with other studies. For example, Feres et al. ^5^ systemically reviewed findings from sequencing studies comparing subgingival plaque in health and periodontitis, as well as periodontitis before and after treatment, and found that common nitrite-producing species were associated with periodontal health (e.g., *Steptococcus* spp., *Neisseria longate*, *Neisseria subflava*, *Rothia aeria*, *Veilonella Parvula* and *Granulicatella adiacens*). Additionally, Feres et al. (2020) found that *Rothia* was the genus with the strongest association with periodontal health (i.e., found significantly higher in healthy individuals after periodontal treatment in most studies) followed by other nitrite-producing genera (e.g., *Neisseria*, *Actinomyces* and *Steptococcus*). These results are further supported by bioinformatic analyses of Meuric et al. ^36^, associating *Veillonella*, *Neisseria*, *Rothia*, *Corynebacterium*, and *Actinomyces* (all genera with nitrite-producing representatives) with periodontal health, and Chen et al. ^6^, who reported that that *Steptococus sanguinis*, *Actinomyces naeslundii*, *Rothia aeria*, *Granulicatella adiacens*, *Rothia dentocariosa* and *Streptococcus mitis* (all nitrite-producing species) were six of the top seven most health-associated species.

The underlying mechanisms by which nitrate reduction by oral bacteria appears to be beneficial for oral health needs to be explored ^7^. Oral bacteria can further reduce nitrite to nitric oxide, a free radical with antimicrobial properties capable of inhibiting sensitive species such as anaerobes associated with periodontitis ^18^. Additionally, nitric oxide derived from denitrification by oral bacteria could directly signal to human cells, reducing inflammation and stimulating protective mucus production and blood flow ^15, 37, 38^. In our study, nitrate-reducing bacteria correlated negatively with periodontitis-associated bacteria, supporting the idea that this function could be important for oral health. This negative correlation was clearest in the 42 periodontitis patients after periodontal treatment. The reduction of inflammation in these patients after treatment was previously described by Johnston et al. ^31^ and consisted of a clear and significant improvement of all periodontal clinical parameters. In our recent *in vitro* study, the addition of nitrate to periodontal plaque resulted in nitrite production (an indication of nitric oxide production), a decrease in periodontitis-associated species and a lower subgingival microbial dysbiosis index ^39^. The impairment of the NRC in periodontitis could thus give a selective advantage to periodontitis-associated species. The effect of nitrate consumption and the patients’ NRC on the efficiency of periodontal treatment should be further investigated in future studies.

One of the main results of this study is that the NRC of saliva samples decreases in periodontitis. Unstimulated saliva contains 10^8^-10^9^ bacteria/ml that have dispersed from oral surfaces ^40^, making this a non-invasive sample that can be used to study the activity of the oral microbiota. Additionally, by freezing at −80°C and thawing on ice, a significant part of the bacteria survives, as demonstrated by the changes in metabolic parameters after 3 hours of incubation at 37°C in this study (e.g., a minor but significant decrease in pH by bacterial fermentation of salivary components). Significant nitrate-reduction activity was only found in the saliva of 15 healthy individuals (p < 0.01) and not in the saliva of 42 periodontitis patients before treatment (p = 0.1). However, 90 days after treatment, the NRC of the individuals with periodontitis was restored (p < 0.001) to levels comparable with healthy individuals. Kapil et al. ^41^ determined the NRC by letting individuals rinse their mouth with a nitrate containing solution. Future studies should compare different methods to determine the NRC, as this could represent a helpful marker of gingival health, and how it correlates with systemic parameters.

In line with the NRC results, we showed that the subgingival nitrate-reducing microbiota decrease in periodontitis (compared with periodontal health) and increases after NSPT. Specifically, we showed this for the relative abundance of known nitrite-producing bacteria, including confirmed nitrate-reducing bacteria. The difference between these two groups is that nitrite-producing isolates are detected by incubating bacteria with nitrate and measuring nitrite production. Most of the time, this nitrite results from nitrate reduction, but there are other pathways that can lead to nitrite production (for example, the oxidation of nitric oxide or ammonium) ^33^. The NRC of oral bacteria should thus be confirmed by physiological measurements of nitrate ^7^. In our study, both groups (nitrite-producing and confirmed nitrate-reducing) showed similar statistical significance (i.e., p < 0.5 remained p < 0.5 when comparing these groups between periodontitis and health or before and after periodontal treatment). Additionally, qPCR measurements of the nitrate-reducing biomarker genus *Rothia* before and after periodontitis treatment confirmed an increase of *Rothia* cells per sample (p < 0.05) and a trend of increase of *Rothia* cells per ng DNA (p = 0.1). Our data thus confirm that nitrate-reducing bacteria decrease in the subgingival plaque under conditions of inflammation and dysbiosis associated with periodontitis. It is also interesting that *Rothia* levels correlated well with the proportion of nitrate-reducing bacteria (Supplementary Datasheet), confirming that this genus is a potential biomarker of nitrate reduction capacity. In fact, the degree of periodontitis, as indicated by the FMB and PISA indexes, showed a (trend of) negative correlation with *Rothia* levels (Supplementary Datasheet). This also supports a possible role of this organism in periodontal health and its assessment as a potential periodontal probiotic, as recently proposed ^31, 39^.

### Implications of hampered NRC for systemic health

Periodontitis increases the risk of systemic diseases ^11^, including comorbidities that are associated with a deficit in nitric oxide ^42, 43^ and/or improve upon nitrate supplementation ^16^, such as cardiovascular disease and diabetes. In this study, we show that the levels of nitrate-reducing bacteria are lower in the subgingival plaque of individuals with periodontitis and increase after non-surgical periodontal therapy (NSPT). The lower abundance of nitrate-reducing bacteria was accompanied by an impaired salivary nitrate reduction capacity (NRC) in individuals with periodontitis that was restored to healthy levels after periodontal treatment. The reduction of nitrate by oral bacteria is an essential step in the nitrate-nitrite-nitric oxide pathway that contributes to systemic nitric oxide levels ^41^. We thus hypothesize that the impairment of NRC in periodontitis can contribute to the development of different comorbidities in addition to known inflammatory and bacterial mechanisms (reviewed by ^11^). Future studies should be designed to determine how periodontitis is affected by the intake of nitrate-rich foods, and what the effect of nitrate-rich diet interventions on systemic nitric oxide levels and associated benefits derived from periodontal health.

In 2006, it was found that sodium nitrate intake reduces blood pressure ^44^. Similarly, beetroot juice, which is particularly high in nitrate, acutely lowers blood pressure in healthy individuals ^45^. In 2013, Kapil et al. ^41^ confirmed that the link between nitrate – an inorganic compound that human cells cannot use effectively – and blood pressure was the NRC of the oral microbiota. When sterilizing a significant proportion of the oral microbiota with chlorhexidine in fasting individuals, the NRC of the oral microbiota was impaired, plasma nitrite levels dropped and blood pressure increased ^41^. Nitrate intake has also been associated with a reversal of metabolic syndrome and with antidiabetic effects^16, 43^, while the use of over-the-counter mouthwash was found to correlate with diabetes and pre-diabetes development ^46^. It thus appears that conditions in which a deficit of nitric oxide is found ^42, 43^, benefit from stimulating nitrate reduction by the oral microbiota. Remarkably, periodontitis is associated with cardiovascular diseases and diabetes ^47, 48^, while in our current study we found a decrease of nitrate-reducing bacteria in subgingival plaque and an impairment of the NRC in patients with periodontitis. We therefore hypothesize that periodontitis contributes to cardiovascular diseases, diabetes, and other nitric oxide-related systemic conditions by stimulating a nitric oxide deficit. This is supported by a recent metagenomic estimation study, associating increased nitrite generation by the oral microbiota with lower levels of cardiometabolic risk ^49^.

Pre-eclampsia is another example of a condition associated with both reduced nitric oxide availability ^50^ and periodontitis ^10^. A recent study indicated that this condition was associated with a decrease in oral nitrate-reducing bacteria ^51^. In relation to this, periodontitis treatment appears to reduce complications in pregnant women ^52^, possibly by reducing inflammatory and bacterial exposure to unborn babies ^10^. In our study, the levels of nitrate-reducing bacteria in subgingival plaque increased after NSPT, while the NRC was restored to healthy levels. Another benefit of periodontal treatment in pregnant women could therefore be to stimulate the NRC and this should be investigated in future studies. Periodontal treatment is also associated with a long-term improvement of endothelial function ^53^. Regarding this, it has been shown that endothelial function can improve when healthy individuals consume nitrate ^16^. In a recent study in mice, it was shown that inorganic nitrate protects against, and can partially reverse pre-existing, periodontitis-induced endothelial function through restoration of nitrite and thus nitric oxide levels ^54^. Altogether, these results indicate that periodontal treatment may decrease the risk of systemic comorbidities of periodontitis by increasing the NRC. Additionally, they indicate that nitrate intake should be tested as adjunct treatment to improve systemic parameters in patients with periodontitis.

Limitations of this study is that individuals were not fasting when donating saliva and plasma samples and did not receive instructions regarding other habits that could interfere with salivary and plasma nitrate and nitrite levels (e.g., exercise, sunlight, water consumption). Considering the large effect of the diet on plasma and salivary nitrate and nitrite levels, the study design was not ideal to see how periodontitis affects the baseline levels of these molecules. This could explain why the plasma levels of nitrate and nitrite were not significantly different before and after periodontal treatment. However, when incubating saliva with nitrate, differences were found in the NRC of periodontitis patients before and after treatment and between periodontitis patients before treatment and healthy individuals. In future studies, this method should be compared with other methods to determine the NRC such as *in vivo* measurements after nitrate-containing mouthwashes used by Kapil et al. ^41^, which take into account the activity of all oral biofilms. Another limitation of the current study is that we did not collect blood samples after conditions of high nitrate availability, which is when the effect of a healthy nitrate-reduction bacterial community is expected to be relevant. Thus, we propose that future studies should test if exposure to nitrate (e.g., by beetroot juice consumption) has different effects on salivary and plasma and nitrate and nitrite levels, as well as their derived systemic effects (e.g., blood pressure), in periodontitis patients and healthy individuals, as well as the potential reversal after periodontal treatment. Finally, in this study the composition of subgingival plaque was studies, which is most effected by periodontitis. However, the effect of this disease on other microbial communities involved in nitrate reduction (e.g., the tongue microbiota) should be explored.

In conclusion, our data show that periodontitis compromises nitrate reduction and that this low nitrate reduction capacity is recovered after non-surgical periodontal treatment. Given that a diminished or eliminated nitrate reduction capacity, for instance by antiseptic mouthwash, derives in lower plasma nitrite levels ^41^, we hypothesize that periodontal disease can imply a deficit in circulatory nitrite. Thus, we propose that, in addition to the oral-systemic link derived from inflammatory effects, a hampered nitrate reduction capacity in periodontal patients could be also responsible for the multiple systemic conditions that are affected by nitric oxide availability. We hope that the current manuscript stimulates further experimental and clinical work to test this hypothesis, including the potential beneficial effects of dietary interventions to improve not only oral but also systemic health mediated by a healthy gingival bacterial community.

## Supporting information

Supplementary Datasheet

## Data Availability

The sequencing reads of the 42 periodontitis patients before (BL) and after (D90) treatment are deposited in the NCBI Sequencing Read Archive (SRA) under BioProject PRJNA725103. Any further data is available upon reasonable request from the corresponding author.

https://www.ncbi.nlm.nih.gov/bioproject/?term=PRJNA725103

## Statement of ethical approval

The study was conducted in accordance with the Declaration of Helsinki (2013). For the study involving periodontitis patients, the London-Stanmore Research Ethics Committee (Reference: 14/LO/206) of National Health Service (NHS, England, United Kingdom) gave ethical approval. For the study involving saliva samples of healthy patients, the MVLS ethical committee (project number: 2011002) of the University of Glasgow (Scotland, United Kingdom) gave ethical approval.

