## Supplementary Figure for "Nitrate reduction capacity of the oral microbiota is impaired in periodontitis: potential implications for systemic nitric oxide availability"

^*^Shared first author

^#^Correspondance:

**Supplementary Information**

*Supplementary Table 1: bacterial species in different groups.*

| **Bacterial group** | **Species^*1^** | **Reference(s)** |
| --- | --- | --- |
| **Nitrite producers^*2^** | *Actinomyces georgiae, Actinomyces graevenitzii, Actinomyces hongkongensis,*  *Actinomyces johnsonii, Actinomyces lingnae, Actinomyces massiliensis, Actinomyces naeslundii, Actinomyces odontolyticus, Actinomyces oris, Actinomyces viscosus, Capnocytophaga gingivalis, Capnocytophaga ochracea, Capnocytophaga sputigena, Corynebacterium durum, Corynebacterium matruchotii, Cutibacterium acnes, Eikenella corrodens, Fusobacterium nucleatum, Granulicatella adiacens, Haemophilus parainfluenzae, Haemophilus segnis, Kingella denitrificans, Neisseria elongate, Neisseria flavescens, Neisseria macacae, Neisseria mucosa, Neisseria oralis, Neisseria sicca, Neisseria subflava, Paraburkholderia fungorum, Prevotella melaninogenica, Prevotella melaninogenica, Propionibacterium acnes, Pseudopropionibacterium propionicum, Rothia aeria, Rothia dentocariosa, Rothia mucilaginosa, Schaalia odontolytica, Selenomonas artemidis, Selenomonas flueggei, Selenomonas noxia, Streptococcus australis, Streptococcus infantis, Streptococcus mitis, Streptococcus mutans, Streptococcus oralis, Streptococcus parasanguinis, Streptococcus salivarius, Streptococcus sanguinis, Veillonella atypica, Veillonella dispar, Veillonella parvula, Veillonella tobetsuensis* | Rosier et al. (2022) |
| **Confirmed nitrate reducers^*3^** | *Actinomyces georgiae, Actinomyces graevenitzii, Actinomyces hongkongensis, Actinomyces johnsonii, Actinomyces lingnae, Actinomyces massiliensis, Actinomyces naeslundii, Actinomyces odontolyticus, Actinomyces oris, Actinomyces viscosus, Cutibacterium acnes, Kingella denitrificans, Neisseria elongate, Neisseria flavescens, Neisseria macacae, Neisseria mucosa, Neisseria oralis, Neisseria sicca,*  *Neisseria subflava, Propionibacterium acnes, Pseudopropionibacterium propionicum, Rothia aeria, Rothia dentocariosa, Rothia mucilaginosa, Schaalia odontolytica, Veillonella atypica, Veillonella dispar, Veillonella parvula, Veillonella tobetsuensis* | Rosier et al. (2022) |
| **Periodontitis associated^*4^** | *Acinetobacter baumannii, Aggregatibacter actinomycetemcomitans, Alloprevotella tannerae, Anaeroglobus geminatus, Campylobacter gracilis, Campylobacter rectus, Campylobacter showae, Dialister pneumosintes, Enterococcus faecalis, Escherichia coli, Eubacterium brachy, Eubacterium nodatum, Eubacterium saphenum, Filifactor alocis, Fretibacterium fastidiuosum, Fusobacterium nucleatum, Fusobacterium nucleatum subsp. animalis, Fusobacterium nucleatum subsp. nucleatum, Fusobacterium nucleatum subsp. polymorphum, Fusobacterium nucleatum subsp. vicentii, Fusobacterium periodonticum, Mogibacterium timidum, Parvimonas micra, Peptostreptococcus stomatis, Porphyromonas endodontalis, Porphyromonas gingivalis, Prevotella denticola, Prevotella intermedia, Prevotella nigrescens, Selenomonas sputigena, Tannerella forsythia, Treponema denticola, Treponema lecithinolyticum, Treponema medium, Treponema vincentii* | Socransky et al. (1998); Pérez-Chaparro et al. (2014) |
| **Red complex^*4^** | *Porphyromonas gingivalis*, *Tannerella forshytia*, *Treponema denticola* | Socransky et al. (1998) |

^*1^In each dataset, the species in these lists that were found were added to these groups
^*2^ Nitrite-producing species were found to produce nitrite in the present of nitrate, but it was not always confirmed if the nitrite production was due to nitrate reduction
^*3^ Confirmed nitrate reducers were considered all nitrite-producing species from genera with confirmed nitrate-reducing representatives
**^*4^** Previously reported and analysed by Johnston et al. (2021)

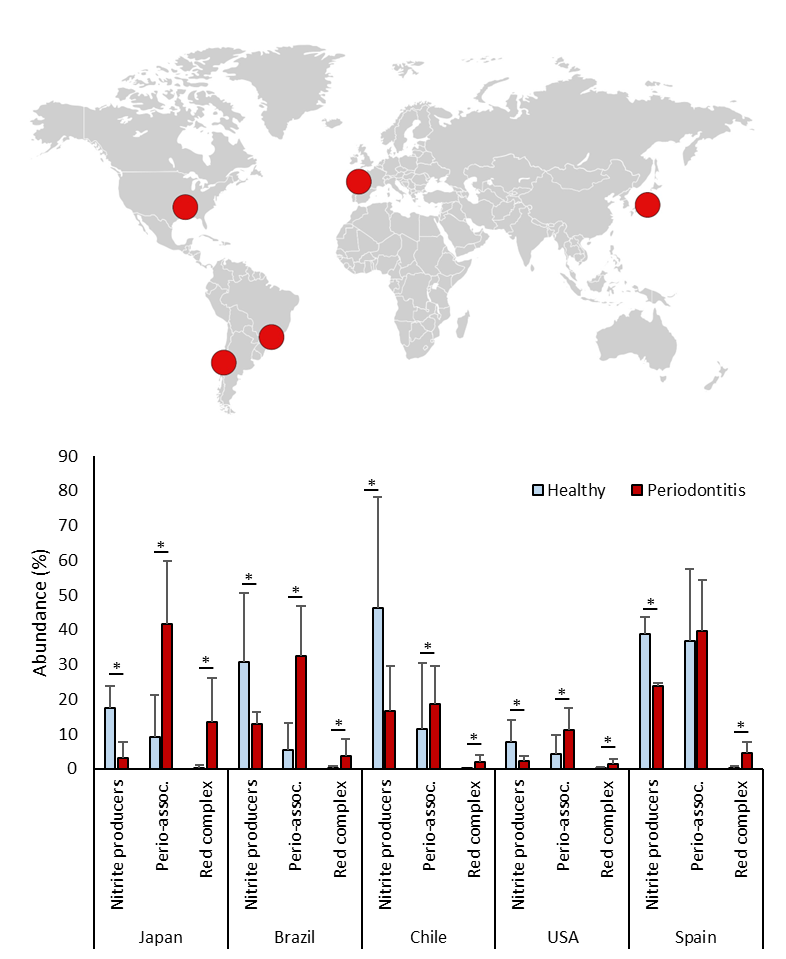

**Supplementary Figure 1: Nitrite-producing bacteria in periodontitis and health.** Bargraphs show the relative abundance of bacteria in subgingival plaque samples from different countries, as estimated by high-throughput sequencing of the 16S rRNA gene. Bacteria were grouped in nitrite-producing species, periodontitis-associated or “red-complex” periodontal pathogens according to Rosier et al. (2022), Pérez-Chaparro et al. (2014) and Socransky et al. (1998), respectively (the bacterial species in each group are listed in Supplementary Table 1). The datasets include individuals from Japan (n = 10 periodontitis patients and 10 healthy individuals), Spain (n = 22 periodontitis patients and 60 healthy individuals), USA (n = 29 periodontitis patients and 28 healthy individuals), Brazil (n = 27 periodontitis patients and 21 healthy individuals) and Chile (n = 22 periodontitis patients and 17 healthy individuals). *adjusted p < 0.05 of compositional data standardized by ANCOM-BC and compared with a Wilcoxon test.

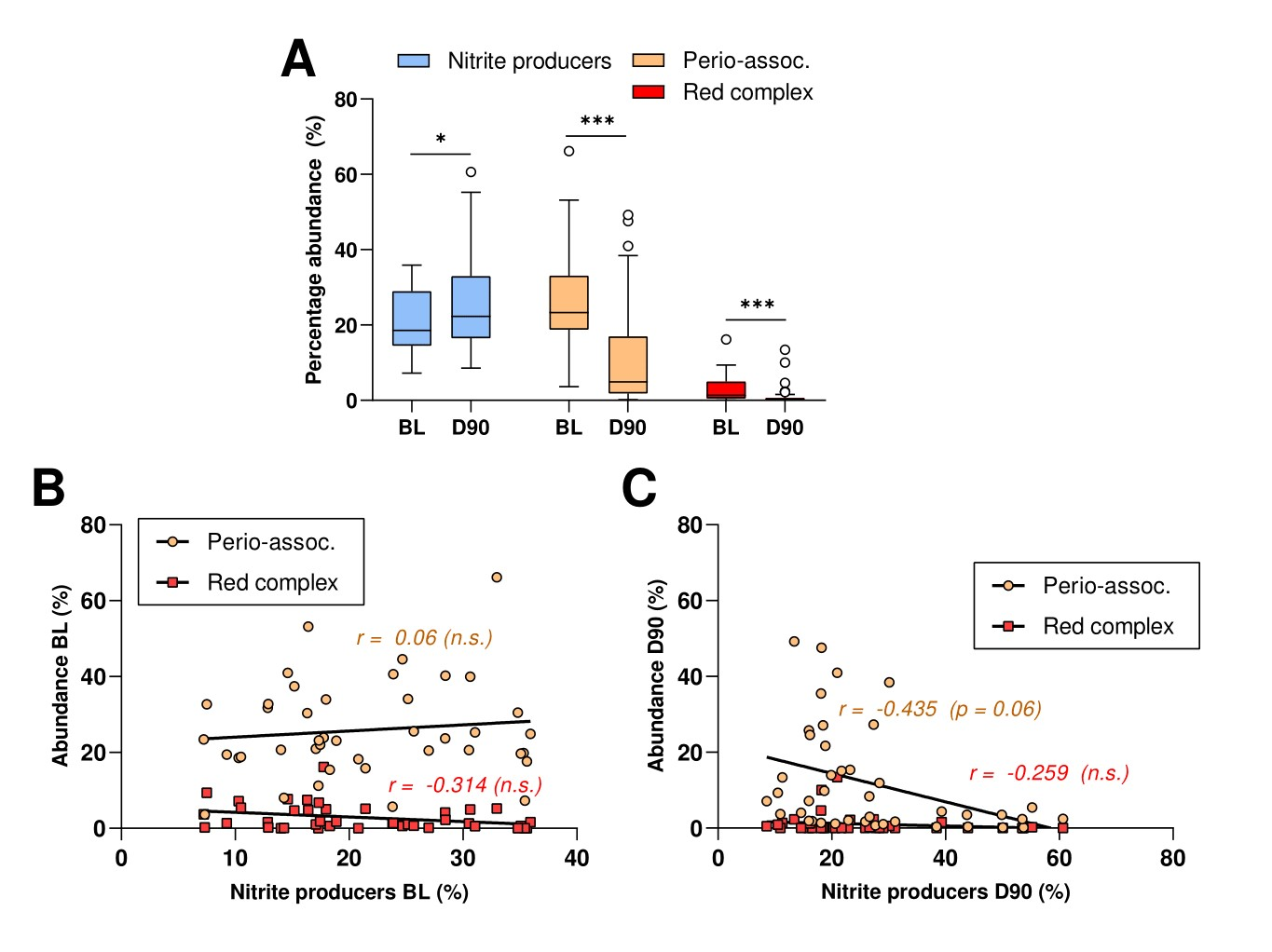

**Supplementary Figure 2: Nitrite-producing species and disease-associated bacteria before and 90 days after periodontal treatment.** A) Relative abundances of nitrite-producing bacteria, red complex and periodontitis-associated bacteria before (baseline, BL) and 90 days after treatment (D90) of 42 periodontitis patients. *adjusted p < 0.05, *** adjusted p < 0.001 of compositional data standardized by ANCOM-BC and compared with a Wilcoxon test. B and C) Correlations between abundance of periodontal pathogens and nitrite-producing bacteria at baseline (BL) and 90 days after treatment (D90). *adjusted p-values of Spearman's rank correlation reported (n.s. = not significant).
