## Supplementary Datasheet for "Nitrate reduction capacity of the oral microbiota is impaired in periodontitis: potential implications for systemic nitric oxide availability"

### Correlations between bacterial groups

BL and D90 (max. N = 84)

| y | x | spearman.cor | cor.adj.p.val | cor.p.val |
|---|---|---|---|---|
| Nitrate_reducers | Perio_assoc | **-0.522790321** | ***8.62E-06*** | 5.15E-07 |
| Nitrate_reducers | Red_compl | **-0.403655494** | ***0.001420211*** | 0.000140389 |
| Nitrite_producers | Nitrate_reducers | **0.593317809** | ***1.04E-07*** | 4.56E-09 |
| Nitrite_producers | Perio_assoc | **-0.251149134** | ***0.097722375*** | 0.021436081 |
| Nitrite_producers | Red_compl | **-0.313274838** | ***0.022745794*** | 0.003712532 |
| Perio_assoc | Red_compl | **0.719443449** | ***4.58E-13*** | 1.26E-14 |
| Rothia_T_ng | Nitrate_reducers | **0.574460454** | ***3.94E-06*** | 1.99E-07 |
| Rothia_T_ng | Nitrite_producers | **0.425705762** | ***0.002209841*** | 0.000238764 |
| Rothia_T_ng | Perio_assoc | **-0.284894453** | ***0.080441156*** | 0.01682792 |
| Rothia_T_ng | Red_compl | **-0.259354513** | ***0.118395385*** | 0.030151759 |
| Rothia_Total | Nitrate_reducers | **0.597201552** | ***1.00E-06*** | 4.84E-08 |
| Rothia_Total | Nitrite_producers | **0.311003397** | ***0.045465444*** | 0.008779534 |
| Rothia_Total | Perio_assoc | **-0.261427973** | ***0.118035477*** | 0.028811902 |
| Rothia_Total | Red_compl | **-0.32253268** | ***0.036535858*** | 0.006467267 |
| Rothia_Total | Rothia_T_ng | **0.788322762** | ***2.17E-14*** | 5.49E-16 |

BL (max. N = 42)

| y | x | spearman.cor | cor.adj.p.val | cor.p.val |
|---|---|---|---|---|
| Nitrate_reducers_BL | Perio_assoc_BL | **-0.320800583** | ***0.221032424*** | 0.038816055 |
| Nitrate_reducers_BL | Red_compl_BL | **-0.390888489** | ***0.0970076*** | 0.010481281 |
| Nitrite_producers_BL | Nitrate_reducers_B | **0.482699943** | ***0.01908762*** | 0.001370069 |
| Nitrite_producers_BL | Perio_assoc_BL | **0.065391783** | ***0.872351649*** | 0.679832664 |
| Nitrite_producers_BL | Red_compl_BL | **-0.313472788** | ***0.241081517*** | 0.04322841 |
| Perio_assoc_BL | Red_compl_BL | **0.431014951** | ***0.048821978*** | 0.004377143 |
| Rothia_T_ng_BL | Nitrate_reducers_B | **0.457595071** | ***0.059152197*** | 0.005711247 |
| Rothia_T_ng_BL | Nitrite_producers_ | **0.334337139** | ***0.258585595*** | 0.049642202 |
| Rothia_T_ng_BL | Perio_assoc_BL | **-0.058407452** | ***0.894173062*** | 0.738923955 |
| Rothia_T_ng_BL | Red_compl_BL | **-0.133520143** | ***0.764212568*** | 0.444473057 |
| Rothia_Total_BL | Nitrate_reducers_B | **0.597479083** | ***0.003646264*** | 0.00015088 |
| Rothia_Total_BL | Nitrite_producers_ | **0.324022935** | ***0.2878775*** | 0.0575755 |
| Rothia_Total_BL | Perio_assoc_BL | **-0.108760968** | ***0.802477321*** | 0.533994567 |
| Rothia_Total_BL | Red_compl_BL | **-0.293313041** | ***0.387283336*** | 0.087250039 |
| Rothia_Total_BL | Rothia_T_ng_BL | **0.840202004** | ***1.99E-08*** | 2.75E-10 |

D90 (max. N = 42)

| y | x | spearman.cor | cor.adj.p.val | cor.p.val |
|---|---|---|---|---|
| Nitrate_reducers_D90 | Perio_assoc_D90 | **-0.583664209** | ***0.0029701*** | 6.83E-05 |
| Nitrate_reducers_D90 | Red_compl_D90 | **-0.33753425** | ***0.245770602*** | 0.028811235 |
| Nitrite_producers_D90 | Nitrate_reducers_D | **0.689490317** | ***6.90E-05*** | 1.11E-06 |
| Nitrite_producers_D90 | Perio_assoc_D90 | **-0.435378008** | ***0.064040795*** | 0.004269386 |
| Nitrite_producers_D90 | Red_compl_D90 | **-0.259178085** | ***0.477018166*** | 0.09743184 |
| Perio_assoc_D90 | Red_compl_D90 | **0.769194979** | ***2.32E-07*** | 2.67E-09 |
| Rothia_T_ng_D90 | Nitrate_reducers_D | **0.615966387** | ***0.004510784*** | 0.000114066 |
| Rothia_T_ng_D90 | Nitrite_producers_ | **0.50952381** | ***0.044016666*** | 0.002023755 |
| Rothia_T_ng_D90 | Perio_assoc_D90 | **-0.420168067** | ***0.143924889*** | 0.012572749 |
| Rothia_T_ng_D90 | Red_compl_D90 | **-0.353407073** | ***0.294918813*** | 0.037288585 |
| Rothia_Total_D90 | Nitrate_reducers_D | **0.564185168** | ***0.012913469*** | 0.000415606 |

| | | | | |
|---|---|---|---|---|
| Rothia_Total_D90 | Nitrite_producers_I | **0.226206318** | ***0.676641438*** | 0.1913262 |
| Rothia_Total_D90 | Perio_assoc_D90 | **-0.318929898** | ***0.390519154*** | 0.061847192 |
| Rothia_Total_D90 | Red_compl_D90 | **-0.312901952** | ***0.416005722*** | 0.067221965 |
| Rothia_Total_D90 | Rothia_T_ng_D90 | **0.676097768** | ***0.000415631*** | 8.29E-06 |

#### Correlations between bacterial groups and clinical parameters

| BL and D90 (max. N = 84) | | | | |
|---|---|---|---|---|
| y | x | **spearman.cor** | ***cor.adj.p.val*** | cor.p.val |
| FMBS | Nitrate_reducers | **-0.226155588** | ***0.135362092*** | 0.038585976 |
| FMBS | Nitrite_producers | **-0.205502988** | ***0.188757788*** | 0.060749633 |
| FMBS | Perio_assoc | **0.382402278** | ***0.002823591*** | 0.000331042 |
| FMBS | Red_compl | **0.399458481** | ***0.001579915*** | 0.000167071 |
| FMBS | Rothia_T_ng | **-0.32124293** | ***0.036870848*** | 0.006696085 |
| FMBS | Rothia_Total | **-0.329203174** | ***0.031685648*** | 0.005390202 |
| FMPS | Nitrate_reducers | **-0.382510697** | ***0.002823591*** | 0.000329644 |
| FMPS | Nitrite_producers | **-0.27425066** | ***0.058606889*** | 0.011586649 |
| FMPS | Perio_assoc | **0.495976616** | ***2.50E-05*** | 1.61E-06 |
| FMPS | Red_compl | **0.44868153** | ***0.000219226*** | 1.86E-05 |
| FMPS | Rothia_T_ng | **-0.258514484** | ***0.118395385*** | 0.030709287 |
| FMPS | Rothia_Total | **-0.252174713** | ***0.130880946*** | 0.035202461 |
| PISA | Nitrate_reducers | **-0.23417148** | ***0.1211786*** | 0.032035722 |
| PISA | Nitrite_producers | **-0.243384851** | ***0.108488236*** | 0.025688019 |
| PISA | Perio_assoc | **0.444327449** | ***0.000256049*** | 2.30E-05 |
| PISA | Red_compl | **0.452383773** | ***0.000188383*** | 1.56E-05 |
| PISA | Rothia_T_ng | **-0.278283235** | ***0.091990085*** | 0.019666846 |
| PISA | Rothia_Total | **-0.281502522** | ***0.086229884*** | 0.018237125 |
| CAL | Nitrate_reducers | **-0.057610335** | ***0.823590277*** | 0.602695269 |
| CAL | Nitrite_producers | **-0.02426924** | ***0.939381976*** | 0.826548289 |
| CAL | Perio_assoc | **0.149351504** | ***0.392663932*** | 0.175119087 |
| CAL | Red_compl | **0.211138799** | ***0.180241675*** | 0.053865328 |
| CAL | Rothia_T_ng | **0.074905515** | ***0.778849928*** | 0.537705114 |
| CAL | Rothia_Total | **0.075558496** | ***0.77837845*** | 0.534155737 |
| PPD | Nitrate_reducers | **-0.137704786** | ***0.434348499*** | 0.211615737 |
| PPD | Nitrite_producers | **-0.159180657** | ***0.355918266*** | 0.148094727 |
| PPD | Perio_assoc | **0.400164032** | ***0.001568718*** | 0.000162281 |
| PPD | Red_compl | **0.37626552** | ***0.003510287*** | 0.000419621 |
| PPD | Rothia_T_ng | **-0.105163695** | ***0.624615633*** | 0.386256564 |
| PPD | Rothia_Total | **-0.119304538** | ***0.571434736*** | 0.325253359 |
| PPD5 | Nitrate_reducers | **-0.236270491** | ***0.118395385*** | 0.030484097 |
| PPD5 | Nitrite_producers | **-0.215306272** | ***0.16589682*** | 0.049196988 |
| PPD5 | Perio_assoc | **0.476512936** | ***6.26E-05*** | 4.61E-06 |
| PPD5 | Red_compl | **0.459709154** | ***0.000135129*** | 1.09E-05 |
| PPD5 | Rothia_T_ng | **-0.145946856** | ***0.45079263*** | 0.227987077 |
| PPD5 | Rothia_Total | **-0.199682524** | ***0.273502505*** | 0.097454916 |

| BL (max. N = 42) | | | | |
|---|---|---|---|---|
| y | x | **spearman.cor** | ***cor.adj.p.val*** | cor.p.val |
| FMBS_BL | Nitrate_reducers_B | **-0.068428745** | ***0.867129913*** | 0.666765767 |
| FMBS_BL | Nitrite_producers_I | **-0.175531081** | ***0.611680606*** | 0.266180668 |
| FMBS_BL | Perio_assoc_BL | **-0.108156333** | ***0.791307394*** | 0.495375455 |

| | | | | |
|---|---|---|---|---|
| FMBS_BL | Red_compl_BL | **0.033336037** | ***0.949706459*** | 0.833995097 |
| FMBS_BL | Rothia_T_ng_BL | **-0.399411329** | ***0.133196238*** | 0.0174533 |
| FMBS_BL | Rothia_Total_BL | **-0.3548557** | ***0.217278574*** | 0.036462841 |
| FMPS_BL | Nitrate_reducers_B | **-0.281451516** | ***0.335567724*** | 0.070970645 |
| FMPS_BL | Nitrite_producers_ | **-0.336428217** | ***0.203724043*** | 0.029372799 |
| FMPS_BL | Perio_assoc_BL | **0.085303081** | ***0.826894118*** | 0.591181772 |
| FMPS_BL | Red_compl_BL | **0.191725824** | ***0.574485428*** | 0.223851503 |
| FMPS_BL | Rothia_T_ng_BL | **-0.307142448** | ***0.340017375*** | 0.07269337 |
| FMPS_BL | Rothia_Total_BL | **-0.25177437** | ***0.503043631*** | 0.144552767 |
| PISA_BL | Nitrate_reducers_B | **-0.098452313** | ***0.802477321*** | 0.533793363 |
| PISA_BL | Nitrite_producers_ | **-0.320314399** | ***0.221032424*** | 0.03912528 |
| PISA_BL | Perio_assoc_BL | **-0.040758447** | ***0.929765522*** | 0.797247218 |
| PISA_BL | Red_compl_BL | **0.169828159** | ***0.612456498*** | 0.28227099 |
| PISA_BL | Rothia_T_ng_BL | **-0.352265566** | ***0.221032424*** | 0.037949902 |
| PISA_BL | Rothia_Total_BL | **-0.281931028** | ***0.434279745*** | 0.100832768 |
| CAL_BL | Nitrate_reducers_B | **-0.212867677** | ***0.528850969*** | 0.175384959 |
| CAL_BL | Nitrite_producers_ | **-0.129892229** | ***0.715075841*** | 0.410963127 |
| CAL_BL | Perio_assoc_BL | **0.074467223** | ***0.85129758*** | 0.638248377 |
| CAL_BL | Red_compl_BL | **0.16780157** | ***0.617439481*** | 0.288138425 |
| CAL_BL | Rothia_T_ng_BL | **-0.12970096** | ***0.774913702*** | 0.457724175 |
| CAL_BL | Rothia_Total_BL | **-0.120343305** | ***0.791307394*** | 0.49106671 |
| PPD_BL | Nitrate_reducers_B | **-0.061193064** | ***0.884001242*** | 0.700260097 |
| PPD_BL | Nitrite_producers_ | **-0.245906961** | ***0.469102374*** | 0.116466796 |
| PPD_BL | Perio_assoc_BL | **0.074323231** | ***0.85129758*** | 0.63994094 |
| PPD_BL | Red_compl_BL | **0.160707048** | ***0.637651283*** | 0.309297519 |
| PPD_BL | Rothia_T_ng_BL | **-0.266377092** | ***0.480051471*** | 0.121917876 |
| PPD_BL | Rothia_Total_BL | **-0.154590291** | ***0.691649751*** | 0.375239865 |
| PPD5_BL | Nitrate_reducers_B | **-0.015976001** | ***0.986068128*** | 0.920012441 |
| PPD5_BL | Nitrite_producers_ | **-0.230557208** | ***0.497594551*** | 0.141843044 |
| PPD5_BL | Perio_assoc_BL | **0.102668106** | ***0.802477321*** | 0.517632774 |
| PPD5_BL | Red_compl_BL | **0.194385867** | ***0.574485428*** | 0.217369228 |
| PPD5_BL | Rothia_T_ng_BL | **-0.228860051** | ***0.539488573*** | 0.186030543 |
| PPD5_BL | Rothia_Total_BL | **-0.08598121** | ***0.839495017*** | 0.623349173 |

| D90 (max. N = 42) | | | | |
|---|---|---|---|---|
| y | x | **spearman.cor** | ***cor.adj.p.val*** | cor.p.val |
| FMBS_D90 | Nitrate_reducers_D | **-0.093792089** | ***0.861014379*** | 0.554652322 |
| FMBS_D90 | Nitrite_producers_ | **-0.002030132** | ***0.996576089*** | 0.989819505 |
| FMBS_D90 | Perio_assoc_D90 | **0.048154726** | ***0.936511215*** | 0.762014902 |
| FMBS_D90 | Red_compl_D90 | **0.14989249** | ***0.794572492*** | 0.343401445 |
| FMBS_D90 | Rothia_T_ng_D90 | **-0.091867235** | ***0.887579612*** | 0.599675102 |
| FMBS_D90 | Rothia_Total_D90 | **-0.053733453** | ***0.936511215*** | 0.759172236 |
| FMPS_D90 | Nitrate_reducers_D | **-0.325206182** | ***0.286774065*** | 0.035599539 |
| FMPS_D90 | Nitrite_producers_ | **-0.100206995** | ***0.861014379*** | 0.527772807 |
| FMPS_D90 | Perio_assoc_D90 | **0.257211476** | ***0.478455557*** | 0.100090703 |
| FMPS_D90 | Red_compl_D90 | **0.093301786** | ***0.861014379*** | 0.55673294 |
| FMPS_D90 | Rothia_T_ng_D90 | **-0.027625881** | ***0.970789295*** | 0.874826216 |
| FMPS_D90 | Rothia_Total_D90 | **0.073837792** | ***0.921104743*** | 0.67335933 |
| PISA_D90 | Nitrate_reducers_D | **-0.062234107** | ***0.928293728*** | 0.695404558 |
| PISA_D90 | Nitrite_producers_ | **-0.015396459** | ***0.973543364*** | 0.922905185 |

| | | | | |
|---|---|---|---|---|
| PISA_D90 | Perio_assoc_D90 | 0.104533852 | 0.861014379 | 0.510010885 |
| PISA_D90 | Red_compl_D90 | 0.192719388 | 0.6814576 | 0.221414903 |
| PISA_D90 | Rothia_T_ng_D90 | -0.020869809 | 0.973543364 | 0.905278331 |
| PISA_D90 | Rothia_Total_D90 | 0.04531447 | 0.951257695 | 0.79603601 |
| CAL_D90 | Nitrate_reducers_D | 0.189951378 | 0.6814576 | 0.228248869 |
| CAL_D90 | Nitrite_producers_I | 0.159157213 | 0.757592612 | 0.314047956 |
| CAL_D90 | Perio_assoc_D90 | -0.114505673 | 0.861014379 | 0.470253415 |
| CAL_D90 | Red_compl_D90 | -0.000371582 | 0.998704843 | 0.998136582 |
| CAL_D90 | Rothia_T_ng_D90 | 0.382406503 | 0.216412145 | 0.023382462 |
| CAL_D90 | Rothia_Total_D90 | 0.475169855 | 0.060893457 | 0.003919579 |
| PPD_D90 | Nitrate_reducers_D | 0.084704552 | 0.887579612 | 0.593796953 |
| PPD_D90 | Nitrite_producers_I | 0.131960775 | 0.835804401 | 0.404818396 |
| PPD_D90 | Perio_assoc_D90 | 0.114776694 | 0.861014379 | 0.469196372 |
| PPD_D90 | Red_compl_D90 | 0.181169635 | 0.6814576 | 0.250880754 |
| PPD_D90 | Rothia_T_ng_D90 | 0.291538257 | 0.4666446 | 0.089269845 |
| PPD_D90 | Rothia_Total_D90 | 0.283642738 | 0.477018166 | 0.098693414 |
| PPD5_D90 | Nitrate_reducers_D | -0.053496232 | 0.930986494 | 0.736513823 |
| PPD5_D90 | Nitrite_producers_I | 0.027073321 | 0.967131058 | 0.864859727 |
| PPD5_D90 | Perio_assoc_D90 | 0.208049936 | 0.674680191 | 0.186118673 |
| PPD5_D90 | Red_compl_D90 | 0.284440653 | 0.416005722 | 0.067899785 |
| PPD5_D90 | Rothia_T_ng_D90 | 0.192869587 | 0.703844066 | 0.266975335 |
| PPD5_D90 | Rothia_Total_D90 | 0.040377424 | 0.951257695 | 0.817862938 |

#### Correlations between bacterial groups and plasma nitrite

| BL and D90 (max. N = 84) | | | | |
|---|---|---|---|---|
| y | x | spearman.cor | cor.adj.p.val | cor.p.val |
| pNO2 | Nitrate_reducers | -0.137765902 | 0.437222579 | 0.217103625 |
| pNO2 | Nitrite_producers | -0.202001513 | 0.209207079 | 0.068769625 |
| pNO2 | Perio_assoc | 0.031007668 | 0.937741483 | 0.78213161 |
| pNO2 | Red_compl | 0.129362567 | 0.471152269 | 0.246733917 |
| pNO2 | Rothia_T_ng | 0.278777699 | 0.097722375 | 0.021330512 |
| pNO2 | Rothia_Total | 0.278293751 | 0.097722375 | 0.021566317 |

| BL (max. N = 42) | | | | |
|---|---|---|---|---|
| y | x | spearman.cor | cor.adj.p.val | cor.p.val |
| pNO2_BL | Nitrate_reducers_B | -0.144425087 | 0.686888064 | 0.366340301 |
| pNO2_BL | Nitrite_producers_I | -0.103310105 | 0.802477321 | 0.519062165 |
| pNO2_BL | Perio_assoc_BL | -0.139198606 | 0.696172769 | 0.384084275 |
| pNO2_BL | Red_compl_BL | 0.149193914 | 0.677251318 | 0.351859305 |
| pNO2_BL | Rothia_T_ng_BL | 0.104515242 | 0.814884098 | 0.556369143 |
| pNO2_BL | Rothia_Total_BL | 0.213270501 | 0.574485428 | 0.225876973 |

| D90 (max. N = 42) | | | | |
|---|---|---|---|---|
| y | x | spearman.cor | cor.adj.p.val | cor.p.val |
| pNO2_D90 | Nitrate_reducers_D | -0.127874564 | 0.835804401 | 0.424239828 |
| pNO2_D90 | Nitrite_producers_I | -0.262020906 | 0.477018166 | 0.098007229 |
| pNO2_D90 | Perio_assoc_D90 | 0.123867596 | 0.835804401 | 0.438999542 |
| pNO2_D90 | Red_compl_D90 | 0.146770682 | 0.812480464 | 0.359824259 |
| pNO2_D90 | Rothia_T_ng_D90 | 0.438349885 | 0.121952951 | 0.010092658 |
| pNO2_D90 | Rothia_Total_D90 | 0.34838414 | 0.331732292 | 0.043468369 |

### Correlations between bacterial groups and salivary IL-1B

BL and D90 (max. N = 84)

| y | x | **spearman.cor** | ***cor.adj.p.val*** | cor.p.val |
|---|---|---|---|---|
| Nitrate_reducers | IL1B | **-0.230482907** | ***0.13280017*** | 0.037448535 |
| Nitrite_producers | IL1B | **-0.335814804** | ***0.014593769*** | 0.002147129 |
| Perio_assoc | IL1B | **0.194762791** | ***0.232347811*** | 0.079585802 |
| Red_compl | IL1B | **0.252328333** | ***0.09953968*** | 0.022196205 |
| Rothia_T_ng | IL1B | **-0.17794192** | ***0.354209306*** | 0.146569368 |
| Rothia_Total | IL1B | **-0.168944691** | ***0.389947687*** | 0.16843577 |

BL (max. N = 42)

| y | x | **spearman.cor** | ***cor.adj.p.val*** | cor.p.val |
|---|---|---|---|---|
| Nitrate_reducers_BL | IL1B_BL | **-0.261498258** | ***0.429334259*** | 0.098697531 |
| Nitrite_producers_BL | IL1B_BL | **-0.546689895** | ***0.005642238*** | 0.000272384 |
| Perio_assoc_BL | IL1B_BL | **-0.019512195** | ***0.982670666*** | 0.903605211 |
| Red_compl_BL | IL1B_BL | **0.149368206** | ***0.677251318*** | 0.35129059 |
| Rothia_T_ng_BL | IL1B_BL | **-0.309114525** | ***0.344618173*** | 0.07526144 |
| Rothia_Total_BL | IL1B_BL | **-0.199083309** | ***0.607592124*** | 0.258995263 |

D90 (max. N = 42)

| y | x | **spearman.cor** | ***cor.adj.p.val*** | cor.p.val |
|---|---|---|---|---|
| Nitrate_reducers_D90 | IL1B_D90 | **-0.133449477** | ***0.835804401*** | 0.404180959 |
| Nitrite_producers_D90 | IL1B_D90 | **-0.009581882** | ***0.984770128*** | 0.952798138 |
| Perio_assoc_D90 | IL1B_D90 | **0.166550523** | ***0.743098277*** | 0.296862682 |
| Red_compl_D90 | IL1B_D90 | **0.160226137** | ***0.757592612*** | 0.316969782 |
| Rothia_T_ng_D90 | IL1B_D90 | **0.008403361** | ***0.986634509*** | 0.962851647 |
| Rothia_Total_D90 | IL1B_D90 | **-0.038658416** | ***0.95368279*** | 0.828157824 |
